# SARS-CoV-2 RNA titers in wastewater anticipated COVID-19 occurrence in a low prevalence area

**DOI:** 10.1101/2020.04.22.20075200

**Authors:** Walter Randazzo, Pilar Truchado, Enric Cuevas-Ferrando, Pedro Simón, Ana Allende, Gloria Sánchez

## Abstract

Severe acute respiratory syndrome coronavirus 2 (SARS-CoV-2) has caused more than 200,000 reported COVID-19 cases in Spain resulting in more than 20,800 deaths as of April 21, 2020. Faecal shedding of SARS-CoV-2 RNA from COVID-19 patients has extensively been reported. Therefore, we investigated the occurrence of SARS-CoV-2 RNA in six wastewater treatments plants (WWTPs) serving the major municipalities within the Region of Murcia (Spain), the area with the lowest COVID-19 prevalence within Iberian Peninsula. Firstly, an aluminum hydroxide adsorption-precipitation concentration method was tested using a porcine coronavirus (Porcine Epidemic Diarrhea Virus, PEDV) and mengovirus (MgV). The procedure resulted in average recoveries of 10.90 ± 3.54% and 10.85 ± 2.11% in influent water and 3.29 ± 1.58% and 6.19 ± 1.00% in effluent water samples for PEDV and MgV, respectively. Then, the method was used to monitor the occurrence of SARS-CoV-2 from March 12 to April 14, 2020 in influent, secondary and tertiary effluent water samples. By using the real-time RT-PCR (RT-qPCR) Diagnostic Panel validated by US CDC that targets three regions of the virus nucleocapsid (N) gene, we estimated quantification of SARS-CoV-2 RNA titers in untreated wastewater waters of 5.38 ± 0.21 log genomic copies/L on average. Two secondary water samples resulted positive (2 out of 18) and all tertiary water samples tested as negative (0 out 12). This environmental surveillance data were compared to declared COVID-19 cases at municipality level, revealing that SARS-CoV-2 was circulating among the population even before the first cases were reported by local or national authorities in many of the cities where wastewaters have been sampled. The detection of SARS-CoV-2 in wastewater in early stages of the spread of COVID-19 highlights the relevance of this strategy as an early indicator of the infection within a specific population. At this point, this environmental surveillance could be implemented by municipalities right away as a tool, designed to help authorities to coordinate the exit strategy to gradually lift its coronavirus lockdown.

## 1. Introduction

Coronaviruses (CoVs) are a family of viruses pathogenic for humans and animals associated to respiratory and gastro-intestinal infections. CoVs used to be considered as minor pathogens for humans as they were responsible of common cold or mild respiratory infections in immunocompetent people. Nonetheless, the emergence of novel and highly pathogenic zoonotic diseases caused by CoVs such as Severe Acute Respiratory Syndrome (SARS), Middle East Respiratory Syndrome (MERS) and most recently SARS-CoV-2 arises questions to be addressed to guide public health response. CoVs are mainly transmitted through respiratory droplets. However, as for SARS and MERS, SARS-CoV-2 RNA has been detected in stool samples from patients suffering COVID-19 and from asymptomatic carriers (He et al., 2020; Pan et al., 2020; Woelfel et al., 2020; Young et al., 2020; Zhang et al., 2020). The duration of viral shedding has been observed to vary among patients with means of 14-21 days (Y. Wu et al., 2020; Xu et al., 2020), as well as the magnitude of shedding varies from 10^2^ up to 10^8^ RNA copies per gram (Lescure et al., 2020; Pan et al., 2020; Woelfel et al., 2020).

Infectious viruses deriving from fecal specimen have been cultured in Vero E6 cells and observed by electron microscopy (Wang et al., 2020). In addition, gastric, duodenal, and rectal epithelial cells are infected by SARS-CoV-2 and the release of the infectious virions to the gastrointestinal tract supports the possible fecal-oral transmission route (Xiao et al., 2020). Even though the possibility of faecal-oral transmission has been hypothesized, the role of secretions in the spreading of the disease is not clarified yet (Wang et al., 2020; Y. Wu et al., 2020; Xu et al., 2020; Yeo et al., 2020).

Wastewater monitoring has been a successful strategy pursued to track chemical and biological markers of human activity including illicit drugs consumption, pharmaceuticals use/abuse, water pollution, and occurrence of antimicrobial resistance genes (Choi et al., 2018; de Oliveira et al., 2020; Lorenzo and Picó, 2019; Mercan et al., 2019). Viral diseases have been also surveilled by the detection of genetic material into wastewater as for enteric viruses (Hellmer et al., 2014; Prevost et al., 2015; Santiso-Bellón et al., 2020), re-emerging zoonotic hepatitis E virus (Cuevas-Ferrando et al., 2020; Miura et al., 2016), and poliovirus during the global eradication programme (Asghar et al., 2014).

Currently, various studies detected SARS-CoV-2 RNA in wastewater worldwide (Ahmed et al., 2020; Lodder and de Roda Husman, 2020; Medema et al., 2020; F. Wu et al., 2020; Wurtzer et al., 2020), and wastewater testing has been suggested as a non-invasive early-warning tool for monitoring the status and trend of COVID-19 infection and as an instrument for tuning public health response (Daughton, 2020; Mallapaty, 2020; Naddeo and Liu, 2020). Under current circumstance, this environmental surveillance could be implemented in wastewater treatment plants as a tool, designed to help authorities to coordinate the exit strategy to gradually lift its coronavirus lockdown. Here, we report the first detection of SARS-CoV-2 RNA in untreated wastewater samples in Spain collected from six different wastewater treatment plants (WWTPs) in Murcia, the lowest prevalence area in Iberian Peninsula. Additionally, the efficacy of the secondary and tertiary treatments implemented in the WWPTs against SARS-CoV-2 has been confirmed. The outcomes of the environmental surveillance reflect the epidemiological data in a low COVID-19 diagnosed cases setting, thus supporting the need of developing and implementing advanced models for wastewater-based epidemiology (WBE).

## 2. Material and methods

### 2.1. Sampling sites and samples collection

Influent and effluent water samples were collected from six WWTPs located in the main cities of the Region of Murcia, Spain (Figure 1). Technical data on WWTPs are provided in Table 1.

**Table 1.** Operating characteristics of WWTPs.

|  | **Population** |  | **Capacity (m<sup>3</sup>/a)** |  | **Reclamation processes** | **Reuse** |
| --- | --- | --- | --- | --- | --- | --- |
|  | Connected | Equivalent | Designed | Current |  |  |
| **Murcia** | 370,893 | 530,499 | 36,500,000 | 36,952,999 | Activated sludge (A2O process), Disinfection, NaClO | Public domain |
| **Cartagena** | 175,870 | 163,969 | 12,775,000 | 8,625,103 | Activated sludge, Disinfection | Irrigation |
| **Molina de Segura** | 67,455 | 150,545 | 9,125,000 | 5,699,930 | Activated sludge, Decantation, Coagulation, Flocculation, Sand filtration, Disinfection, UV, NaClO | Irrigation |
| **Lorca** | 73,057 | 101,161 | 7,300,000 | 3,366,919 | Activated sludge, Coagulation, Flocculation, Sand filtration, Disinfection, UV, NaClO | Irrigation |
| **Cieza** | 33,744 | 69,502 | 3,650,000 | 2,338,673 | Activated sludge (Extended aeration), Disinfection, Coagulation, Flocculation, Sand filtration, Disinfection, UV | Irrigation |
| **Totana** | 29,113 | 28,289 | 2,190,000 | 1,440,463 | Activated sludge (Extended aeration), Disinfection, UV | Irrigation |

**Figure 1.**
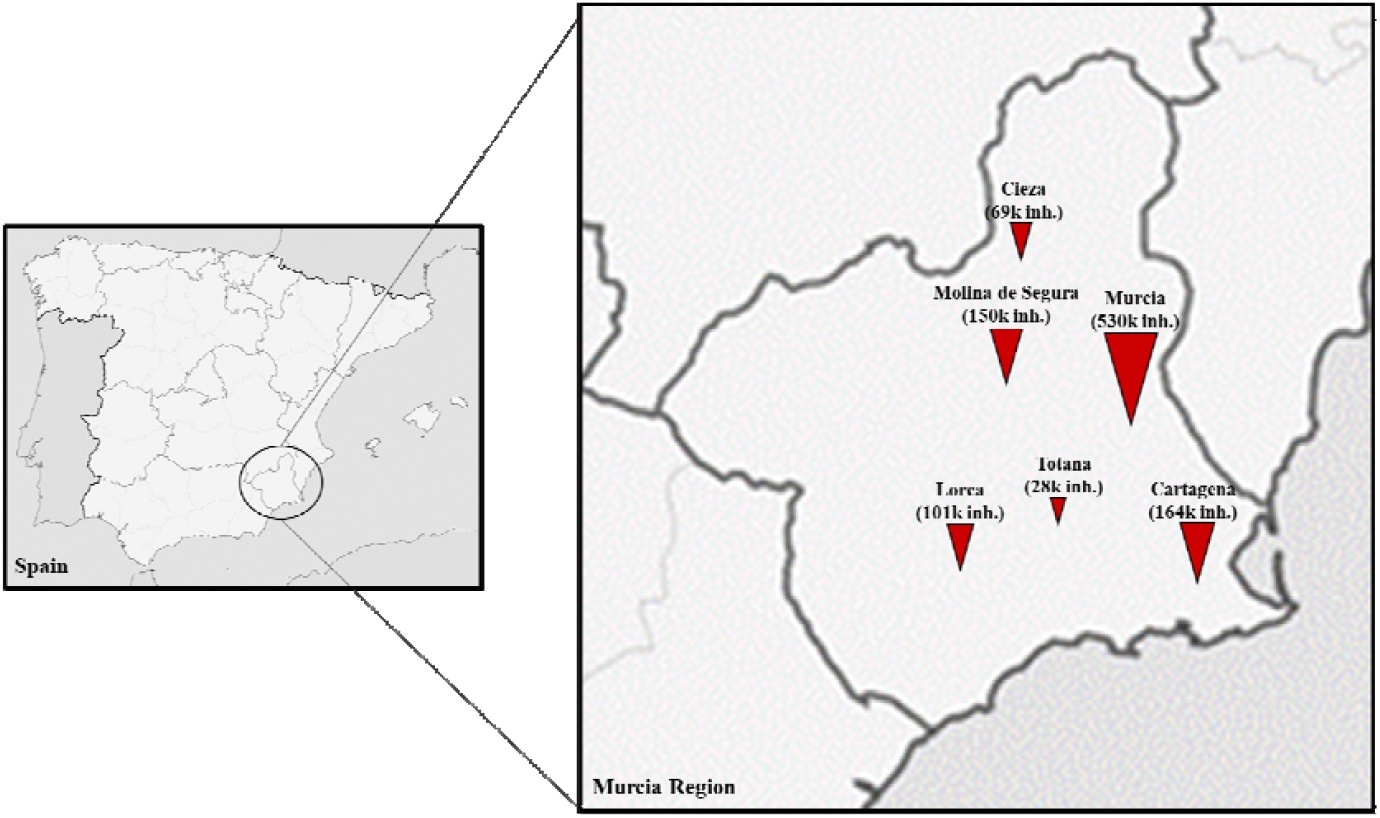
Maps of the sampling location. Symbols represents WWTPs and are sized according to the number of equivalent inhabitants.

A total of 42 influent, and 18 secondary and 12 tertiary treated effluent water samples were collected from 12 March to 14 April 2020 and investigated for the occurrence of SARS-CoV-2 RNA. Collected samples were transferred on ice to the laboratory and concentrated on the same day of sampling or the day after.

### 2.2. Wastewater and effluent water concentration

The porcine epidemic diarrhea virus (PEDV) strain CV777 and the mengovirus (MgV) vMC0 (CECT 100000) were preliminary used to evaluate the aluminum hydroxide adsorption-precipitation method previously described for concentrating enteric viruses from wastewater and effluent water (AAVV, 2011; Randazzo et al., 2019). In brief, 200 mL of biobanked influent (n=2) and effluent water samples (n=2) were artificially inoculated with PEDV and MgV. Then pH was adjusted to 6.0 and Al(OH)^3^ precipitate formed by adding 1 part 0.9N AlCl^3^ (Acros organics, Geel, Belgium) solution to 100 parts of sample. The pH was readjusted to 6.0 and sample mixed using an orbital shaker at 150 rpm for 15 min at room temperature. Then, viruses were concentrated by centrifugation at 1,700 × g for 20 min. The pellet was resuspended in 10 mL of 3% beef extract pH 7.4, and samples were shaken for 10 min at 150 rpm. Concentrate was recovered by centrifugation at 1,900 × g for 30 min and pellet resuspended in 1 mL of PBS.

All wastewater and effluent water samples included in this study were processed as described and MgV (5 log PCRU) was spiked as process control.

### 2.3. Viral extraction, detection and quantification

Viral RNA was extracted from concentrates using the NucleoSpin RNA virus kit (Macherey-Nagel GmbH & Co., Düren, Germany) according to the manufacturer’s instructions with some modifications. Briefly, 150 µL of the concentrated sample was mixed with 25 µL of Plant RNA Isolation Aid (Thermo Fisher Scientific, Vilnius, Lithuania) and 600 µL of lysis buffer from the NucleoSpin virus kit and subjected to pulse-vortexing for 1 min. Then, the homogenate was centrifuged for 5 min at 10,000 × g to remove the debris. The supernatant was subsequently processed according to the manufacturer’s instructions.

Viral RNA was detected by TaqMan real-time RT-PCR (RT-qPCR) on LightCycler 480 instrument (Roche Diagnostics, Germany). MgV RNA was detected by using UltraSense One-Step kit (Invitrogen, SA, US) and the RT-qPCR assay as in ISO 15216-1:2017 (Costafreda et al., 2006; ISO 15216-1, 2017). Undiluted and ten-fold diluted MgV RNA was tested to check for RT-qPCR inhibitors.

PEDV RNA was detected by using PrimeScript™ One Step RT-PCR Kit (Takara Bio, USA) and the TaqMan RT-qPCR assay described by (Zhou et al., 2017).

SARS-CoV-2 RNA was detected by using PrimeScript™ One Step RT-PCR Kit and the RT-qPCR diagnostic panel assays validated by the US Centers for Disease Control and Prevention (CDC, 2020). The first version of the kit with three sets of oligonucleotide primers and probes was used to target three different SARS-CoV-2 regions of the nucleocapsid (N) gene. The sets of primer and probes (2019-nCoV RUO Kit) as well as the positive control (2019-nCoV_N_Positive Control) were provided by IDT (Integrated DNA Technologies, Leuven, Belgium). Each RNA was analyzed in duplicate and every RT-qPCR assay included negative (nuclease-free water) and positive controls.

Biobanked samples (n=4) collected in October 2019, before the first COVID-19 case was documented, were used as relevant negative control to exclude false positive reactions.

SARS-CoV-2 RNA was quantified as genome copies (gc) by plotting the quantification cycles (Ct) to an external standard curve built with 10-fold serial dilution of a quantified plasmid control (IDT). MgV and PEDV RNA were quantified by plotting the Cts to an external standard curve generated by serial end-point dilution method.

MgV recovery rates were calculated and used as quality assurance parameters according to ISO 15216-1:2017 (ISO 15216-1, 2017).

## 3. Results and Discussion

### 3.1. Performance of the concentration method

The aluminum hydroxide adsorption-precipitation method was tested by spiking influent and effluent samples with MgV and PEDV. On average, MgV was recovered at ranges of 10.85 ± 2.11% in influent and 6.19 ± 1.00% in effluent water. PEDV was recovered at ranges of 10.90 ± 3.54% in influent and 3.29 ± 1.58% in effluent water. These results are in line with the MgV recoveries reported for enteric viruses concentration in water samples by the same aluminum-based method (Cuevas-Ferrando et al., 2020; Randazzo et al., 2019) and higher than the 1% as the quality assurance parameter indicated for bottled water into ISO 15216-1:2017 (ISO 15216-1, 2017).

Similarly, MgV was successfully used as recovery control for hepatitis E virus concentration from influent and effluent water samples (5-13%) by applying a polyethylene glycol (PEG) precipitation method (Miura et al., 2016). A similar PEG-based protocol was recently used to recover SARS-CoV-2 from wastewater, although recovery control was not included in the study (F. Wu et al., 2020).

Moreover, filtration through 10 kDa Centricon^®^ Plus-70 centrifugal device successfully recovered SARS-CoV-2 in wastewater with recovery efficiencies of F-specific RNA phages of 73% (Medema et al., 2020). However, concentration by electropositive membrane should be further evaluated given a SARS-CoV recovery from wastewater of 1% (Wang et al., 2005).

Rigorous limits of detection should be established by spiking SARS-CoV-2 cell-culture adapted strain or positive COVID-19 fecal samples in influent and effluent wastewater samples to be concentrated following the aluminum hydroxide adsorption-precipitation method. Nonetheless, the need of a BSL3 laboratory facility to handle SARS-CoV-2 represents the main limitation of this experiment.

### 3.2. SARS-CoV-2 titers in wastewater and effluent water

A total of 42 influent, and 18 secondary and 12 tertiary treated effluent water samples were collected from 12 March to 14 April 2020 and investigated for the occurrence of SARS-CoV-2 RNA. Samples were considered positive for Ct below 40 (as in Medema et al., 2020 and F. Wu et al., 2020) and titrated by using the quantified plasmid control for each of the RT-qPCR targets.

The 83.3% (35 positive samples out of 42) influent samples and the 11.1% (2 out of 18) secondary treated water samples were tested positive for at least one SARS-CoV-2 RT-qPCR target. None of the tertiary effluent samples (0 out of 12) tested positive for any of the SARS-CoV-2 RT-qPCR target (Figure 2). A relevant number of influent water samples (12%) showed Ct ranging between 37 and 40, even though lower Ct of 34-37 were observed (29%). In all samples, MgV recoveries were above 1% (10.05 ± 14.10%).

**Figure 2.**
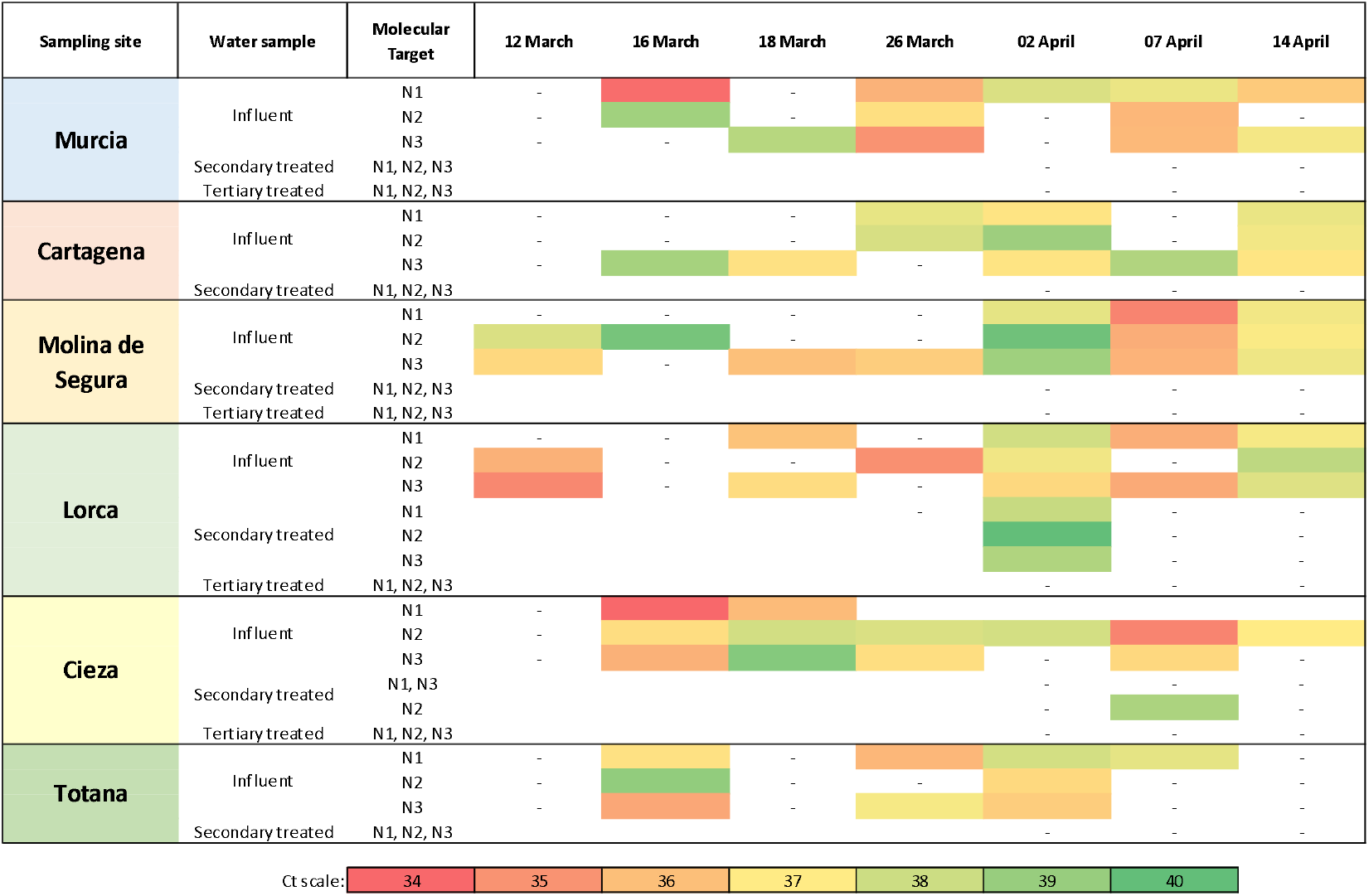
Mean amplification cycles of SARS-CoV-2 RNA in influent, secondary and tertiary effluent waters in monitored WWTPs within Murcia Region (Spain). Results are reported for each of the three regions of the virus nucleocapsid (N) gene according to the first version of the Real-Time RT-PCR Diagnostic Panel by US CDC. Abbreviations: -, negative; white boxes, not tested.

On average, SARS-CoV-2 RNA titers of 5.15 ± 0.25, 5.53 ± 0.24, and 5.49 ± 0.27 log gc/L were quantified in wastewater by using N1, N2 and N3 primer/probe mixes, respectively. Titers of 4 and 5 to more than 6 log gc/L have been reported in Massachusetts and France, respectively (F. Wu et al., 2020; Wurtzer et al., 2020).

A secondary effluent sample resulted positive for N2 and quantified as 5.40 log gc/L. An additional secondary effluent sample was positive for the three molecular targets and below the quantification limit.

Detection of SARS-CoV-2 RNA in influent water has been reported worldwide (Ahmed et al., 2020; Lodder and de Roda Husman, 2020; Medema et al., 2020; F. Wu et al., 2020), and only one study tested treated wastewater that resulted positive (Paris) (Wurtzer et al., 2020). We observed discrepancies among RT-qPCR N1, N2 and N3 assays for several water samples in agreement to a previous report (Medema et al., 2020). This could be due to the different analytical sensitivity among the assays as well as the detection of possible false positive samples by RT-qPCR N3 in low concentrated samples (Jung et al., 2020; Vogels et al., 2020). The latter possibility has been solved by excluding the N3 primers/probe set from the US CDC 2019-nCoV RT-qPCR diagnostic panel in its last revision (March, 30) (CDC, n.d.). In addition, a partial inhibitory effect of the matrix is not to be completely excluded despite the controls included in the assays. A more sensitive estimation of SARS-CoV-2 loads in wastewater should be studied by digital RT-qPCR (dRT-qPCR). dRT-qPCR could be used to quantify samples with low viral loads, even though it may not be the best practical and economically sustainable option for environmental surveillance.

Even though the SARS-CoV-2 RNA detection in wastewater is functional for WBE purposes, the risk for human health associated to the water cycle is still under debate as infectivity of viral particles remain to be confirmed as well as its potential fecal-oral transmission.

In spite of the high concentration of viral RNA in specimen and the evidence of gastrointestinal infection (Xiao et al., 2020), infectious viruses from stools have been isolated in one study (Wang et al., 2020) while another attempt resulted without success (Woelfel et al., 2020).

The potential transmission of SARS-CoV-2 via wastewater has not been proven (CDC, n.d.; WHO, 2020) and it seems unlikely given the poor viral stability in environmental conditions and its elevated sensitivity to disinfectants (Chin et al., 2020; Haas, 2020; Lodder and de Roda Husman, 2020).

### 3.3. Environmental surveillance

Epidemiological data on COVID-19 in the Murcia Region have been retrieved from the publically available repository of the “Servicio de epidemiologia” of the “Consejería de Salud de la Región de Murcia” (available at http://www.murciasalud.es/principal.php) (Table 2) and plotted to the SARS-CoV-2 RNA mean loads as detected by three RT-qPCR assays (Figure 3).

**Table 2.** Epidemiological data^1^ summary of COVID-19 cases in the area of study.

|  | 20/03/2020 |  | 25/03/2020 |  | 30/03/2020 |  | 08/04/2020 |  | 15/04/2020 |  |
| --- | --- | --- | --- | --- | --- | --- | --- | --- | --- | --- |
|  | Cases | Prevalence<sup>2</sup> | Cases | Prevalence | Cases | Prevalence | Cases | Prevalence | Cases | Prevalence |
| **Murcia** | 96 | 21.18 | 210 | 46.33 | 332 | 73.2 | 551 | 121.6 | 622 | 137.2 |
| **Cartagena** | 36 | 16.76 | 64 | 29.79 | 111 | 51.7 | 163 | 75.9 | 190 | 88.5 |
| **Molina de Segura** | 12 | 16.69 | 26 | 36.17 | 40 | 55.6 | 60 | 83.5 | 70 | 97.4 |
| **Lorca** | - | - | 8 | 8.47 | 18 | 19.1 | 29 | 30.7 | 31 | 32.8 |
| **Cieza** | - | - | 12 | 34.30 | 22 | 62.9 | 45 | 128.6 | 49 | 140.0 |
| **Totana** | - | - | - | - | 7 | 21.9 | 13 | 40.6 | 14 | 43.7 |

<sup>1</sup> Data retrieved from the public repository of the “Servicio de epidemiología” of the “Consejería de Salud de la Región de Murcia” (available at http://www.murciasalud.es/principal.php).
<sup>2</sup> Prevalence, percentage of diagnosed cases per 100.000 inhabitants.

**Figure 3.**
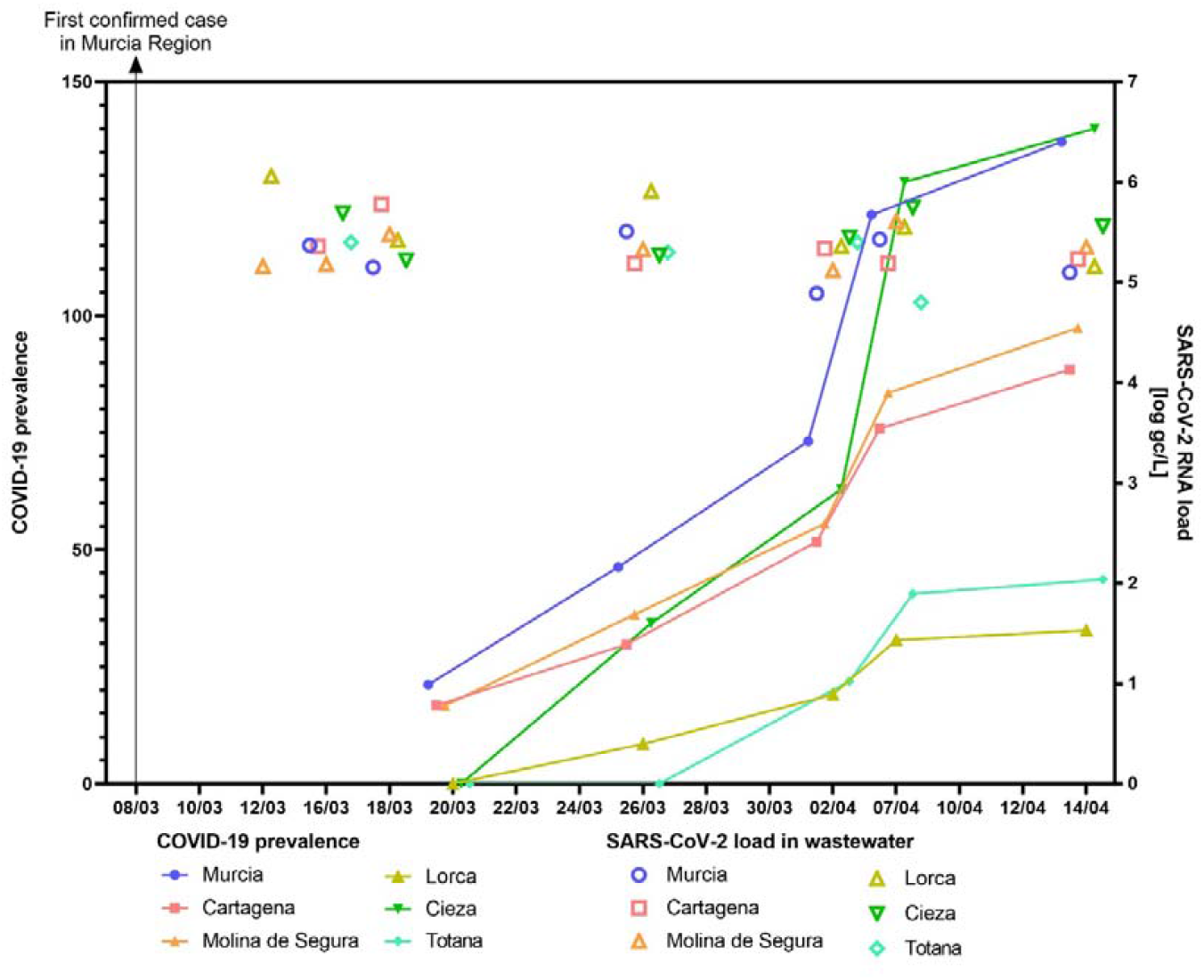
Epidemiological surveillance of COVID-19 by wastewater SARS-CoV-2 RT-qPCR in six municipalities.

In general, RT-qPCR amplification signals have been detected in wastewaters when cases were diagnosed within the municipality. Positive wastewater samples have been detected with at least two out of three RT-qPCR assays in low prevalence municipalities as in Murcia (96 cases, 21.18 cases per 100,000 inhabitants), Cartagena (36 cases, 16.76) and Molina de Segura (12 cases, 16.69). Of note, positive wastewater samples were detected 12-16 days before COVID-19 cases were declared in Lorca, Cieza and Totana municipalities.

A similar study conducted in Paris (France) demonstrated the detection of viral genome before the exponential phase of the epidemic (Wurtzer et al., 2020). However, our results indicate that SARS-CoV-2 can be detected weeks before the first confirmed case. The early detection of SARS-CoV-2 RNA in wastewater could have alerted about the imminent danger, giving a valuate time to the managers to coordinate and implement actions to slow the spread of the disease. Therefore, our outcomes support that WBE could be used as an early warning tool to monitor the status and the trend of COVID-19 infection within a community.

On the other hand, we believe that this environmental surveillance could be used as an instrument to drive the right decisions to reduce the risk of lifting restrictions too early. For instance, a key question is how to reduce the risk of a “second wave” and/or recurring local outbreaks. Massive population tests are the first choice, but in their absence, wastewater monitorization of SARS-CoV-2 RNA can give a reliable picture of the current situation. Our wastewater data do not quantitatively resemble the prevalence of COVID-19 confirmed cases. To this end, a quantitative model that includes and corrects all the variables affecting these wastewater surveillance data would be useful for a better interpretation. For instance, not all COVID-19 positive patients excrete SARS-CoV-2 RNA in faeces, and when it occurs, the titers and the duration of shedding vary among individuals and across time (He et al., 2020; Pan et al., 2020; Woelfel et al., 2020; Xu et al., 2020). On the other hand, the real number of positive cases within the Murcia Region remains unknown because of the large number of mild or asymptomatic carriers that have not been included in epidemiological statistics.

These aspects together with environmental variables (e.g., rainfall events, temperature) increase the uncertainties linked to the correlation between SARS-CoV-2 RNA detection in wastewater samples and the prevalence of COVID-19 that could be explored by using complex models.

## 4. Conclusion

Overall, wastewater surveillance and WBE may represent a complementary approach to estimate the presence and even the prevalence of COVID-19 in communities. This represents an effective tool that needs to be further explored in order to direct public health response, especially in cases of limited capacity for clinical testing.

## Data Availability

Full data set are available under request.

## Acknowledgments

The authors acknowledge the “Entidad Regional de Saneamiento y Depuración de Aguas Residuales (ESAMUR)” for authorizing the sampling and Prof. Ana Carvajal (Faculty of Veterinary Medicine, University of Leon, Spain) for kindly providing PEDV.

